# Comparing supervised machine learning algorithms for the prediction of partial arterial pressure of oxygen during craniotomy

**DOI:** 10.1101/2022.06.07.22275483

**Authors:** Andrea S Becker-Pennrich, Maximilian M Mandl, Clemens Rieder, Dominik J Hoechter, Konstantin Dietz, Benjamin P Geisler, Anne-Laure Boulesteix, Roland Tomasi, Ludwig C Hinske

**Author notes:** Address correspondence to Prof. Hinske: Piechlerstraße 3-5, 86356 Neusäß, Germany.

## Abstract

**Objective:** To evaluate the feasibility of continuous paO_2_ prediction in an intraoperative setting among neurosurgical patients with modern machine learning methods.

**Materials and Methods:** Data were extracted from routine clinical care of lung-healthy, neurosurgical patients. We used recursive feature elimination to identify relevant features for the prediction of paO_2_. Five machine learning algorithms (gradient boosting regressor, k-nearest neighbors regressor, random forest regressor, support vector regression, multi-layer perceptron regressor) and a multivariate linear regression were then tuned and fitted to the selected features. A performance matrix consisting of Spearman’s ρ, mean absolute percentage error (MAPE) and root mean squared error (RMSE) was finally computed based on the test set and used to compare and rank each algorithm.

**Results:** We analyzed 4,180 patients with 12,497 observations. A total of 20 features were selected from analysis of the training dataset comprising 836 patients with 9,992 observations. The best algorithm, random forest, was able to predict paO_2_ values with ρ=0.90, MAPE=10.4%, and RMSE=30.9mmHg, closely followed by gradient boosting and multi-layer perceptron. Support vector regression, k-nearest neighbors regressor and the linear regression did not achieve the performance metrics.

**Discussion:** We successfully applied and compared several machine learning algorithms to estimate continuous paO_2_ values in neurosurgical patients. The random forest regressor performed best over all three categories of the performance matrix.

**Conclusion:** PaO_2_ can be predicted by perioperative routine data in neurosurgical patients.

## INTRODUCTION

Brain tissue is exquisitely sensitive to reduced oxygen levels in the blood, hypoxemia. Hypoxemia leads to irreversible brain damage including brain death within minutes [1]. Consequently, hypoxemia has been the subject of extensive research since the mid 19^th^ century [2]. On the other hand, hyperoxemia, an increased amount of oxygen in the blood, may damage lipids, proteins, and/or nucleic acids, and may even lead to acute lung injury [2]. Diagnosing hyperoxemia is more difficult as its initial symptoms - if any - may be vague [2, 3] and confirmation requires an arterial blood draw, which is different from hypoxemia where peripheral pulse oximetry is often sufficient. Consequently, hyperoxemia has been studied less than hypoxemia and there still is no consensus regarding the potential over-supplementation of oxygen [3–7].

Arterial blood gas analysis (ABG) is performed during surgeries and in the intensive care unit and allows one to draw conclusions about gas exchange in the lungs as well as about tissue oxygenation and oxygen consumption. However, these measurements are only valid at the time of each arterial blood draw, which due to their invasive nature are only infrequently taken. To overcome this limitation, several approaches have been developed aiming at a non-invasive and continuous estimation of the partial oxygen pressure (paO_2_) as a proxy for blood oxygenation [11–17]. These methods estimate blood oxygenation from the fraction of inspired oxygen (FiO_2_) and oxygen saturation (SpO_2_) [8–10], from the oxygen transfer slope and estimated membrane oxygen transfer [11], via the alveolar gas equation [12], and from venous blood gas samples [13–16]. However, we will show that none of these methods are particularly precise and that they also face other limitations such as upper or lower limits of utilized formulas.

Machine learning algorithms may be able to overcome these limitations and give a potentially better accuracy when predicting outcomes based on a higher number of features, non-linear effects, and complex association patterns [18–20].

We therefore attempted to demonstrate that machine learning algorithms are able to predict paO_2_ values better than surrogate parameters or existing equations, i.e., with a satisfiable range of error and good performance parameters. Our second aim was to identify the most accurate machine learning algorithm to predict paO_2_ values nearly continuously.

## MATERIALS AND METHODS

### Data and Data Preparation

The study was conducted as a single center retrospective cohort study. The University of Munich’s institutional review board approved our protocol (submission 19-539) prior to data access. Due to the retrospective design and anonymization, written consent was not necessary.

All patients requiring anesthesia for intracranial surgical procedures at the University Hospital of Munich between January 1^st^ 2009 and December 31^st^ 2019 were screened for inclusion in this study.

Inclusion criteria were: age ≥18 years, intervention type “craniotomy” (identified by the German surgical procedure classification [21] codes 5-01 or 5-02 without 5-023, 5-024 and 5-029), general anesthesia with endotracheal intubation, documented anesthesia induction, incision, closure, and termination of anesthesia times, at least two intraoperatively measured paO_2_ values. In case of multiple surgeries per patient, only the first surgical intervention was included.

Data were extracted and integrated from the anesthesia information management system (AIMS, NarkoData^Ⓡ^, IMESO IT GmbH, Giessen, Germany) and the Hospital Information System (SAP/Cerner i.s.h.med, Idstein, Germany) prior to data anonymization.

Variables known to affect oxygenation were pre-selected (FiO_2_, end-tidal CO_2_, SpO_2_, respiratory minute volume, respiratory rate, blood pressure and heart rate, hemoglobin, paO_2_). We also included the underlying physiologic model of the alveolar gas equation assuming a prevailing atmospheric pressure of 713.622 mmHg and a saturated vapor pressure of water of 47 mmHg [12] as well as the intraoperatively measured paO_2_/FiO_2_ ratio (Horowitz index) as a measure of pulmonary function. The ventilation compliance (static compliance 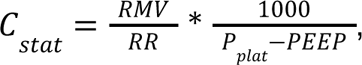, where *RMV* is the respiratory minute volume, *RF* is the respiratory rate, *P_plat_* is the plateau pressure and *PEEP* is the positive end-expiratory pressure) was included as well [22]. Finally, a paO_2_ value based on Gadrey et al. with 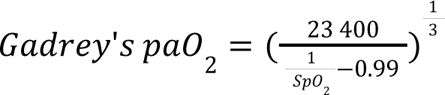 was calculated [17]. One observation was defined as a set of multiple measurements like the aforementioned features as well as one time measurements such as socio-demographic information. For the full set of variables see S1 Appendix. Throughout each surgical procedure, many observations were collected.

Exclusion criteria were missing length of stay, patient height ≤ 120 cm, less than two documented intraoperative paO_2_ value measurements, or an initial intraoperative paO_2_/FiO_2_ ratio <300 mmHg, representing an impaired oxygenation capacity of the lungs. Observations were ignored when they had missing FiO_2_, CO_2_, and SpO_2_ values. We also removed observations with body temperature measurements ≤32°C as the temperature sensor was likely disconnected. Finally, observations with measured paO_2_ values <60 mmHg were excluded in the presence of hemoglobin values >7 g/dL and peripheral SpO_2_ values >95 %, since these values most likely represent a misclassification of a venous blood gas analysis as arterial (see Figure 1).

**Figure 1.**
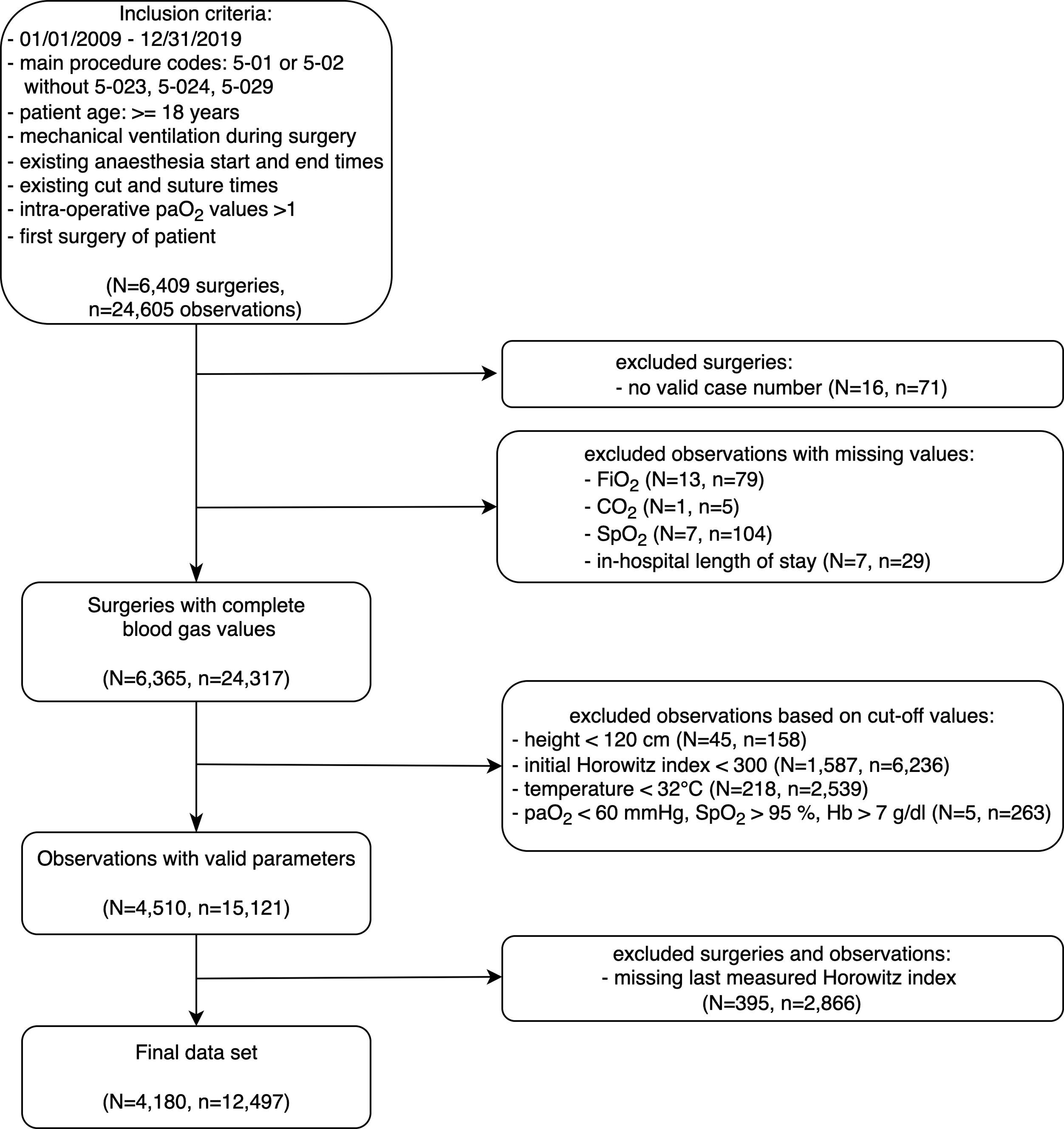
Patient Flowchart.

Remaining missing data (preoperative creatinine, respiratory rate, compliance, respiratory minute volume, blood pressure and heart rate, blood pH values, and hemoglobin) were imputed in a round-robin fashion by a trained iterative imputer with ten iterations using a Bayesian Ridge regression. The completed dataset was used for further analyses.

### Training and Test Datasets

The dataset was randomly divided by a 4:1 ratio into a training and test set, such that the training and test set did not have overlapping patients to impede an overoptimistic bias in the performance evaluation.

Only the training set was used in the feature selection process as well as for performing hyperparameter tuning of the machine learning algorithms (see Figure 2). The test set was used for the purpose of evaluation only.

**Figure 2.**
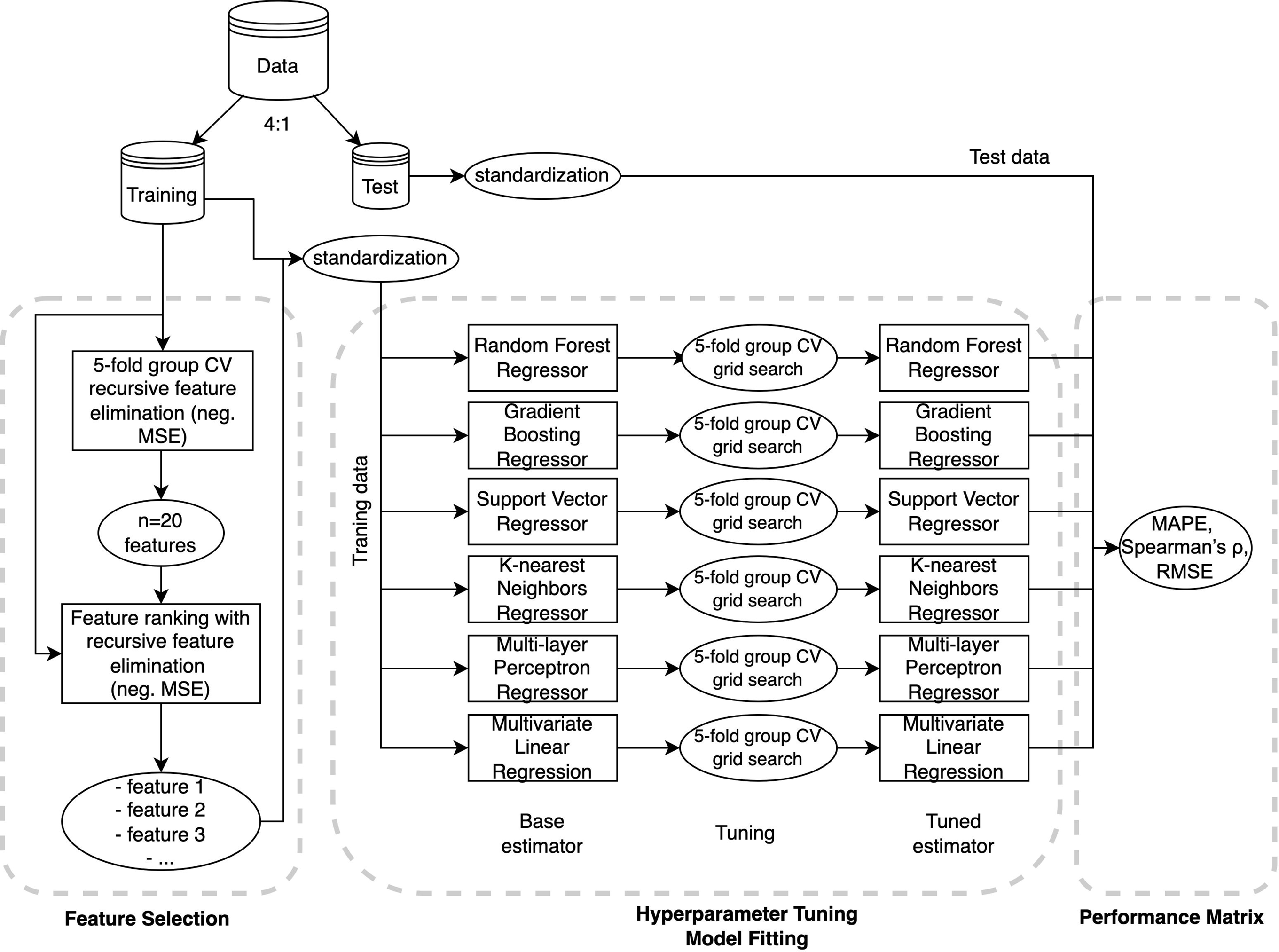
Methods Flowchart.

### Feature Selection

Feature selection was conducted using the recursive feature elimination (RFE) method with random forest regressor as estimator. The optimal number of features was determined using a 5-fold group cross-validation, where each patient was randomly assigned to one of the five folds with the negative mean squared error (neg. MSE) as a performance metric. The optimal number of features was selected as a compromise between computational costs (number of features) and performance (cross-validation scores) (see Figure 3).

**Figure 3.**
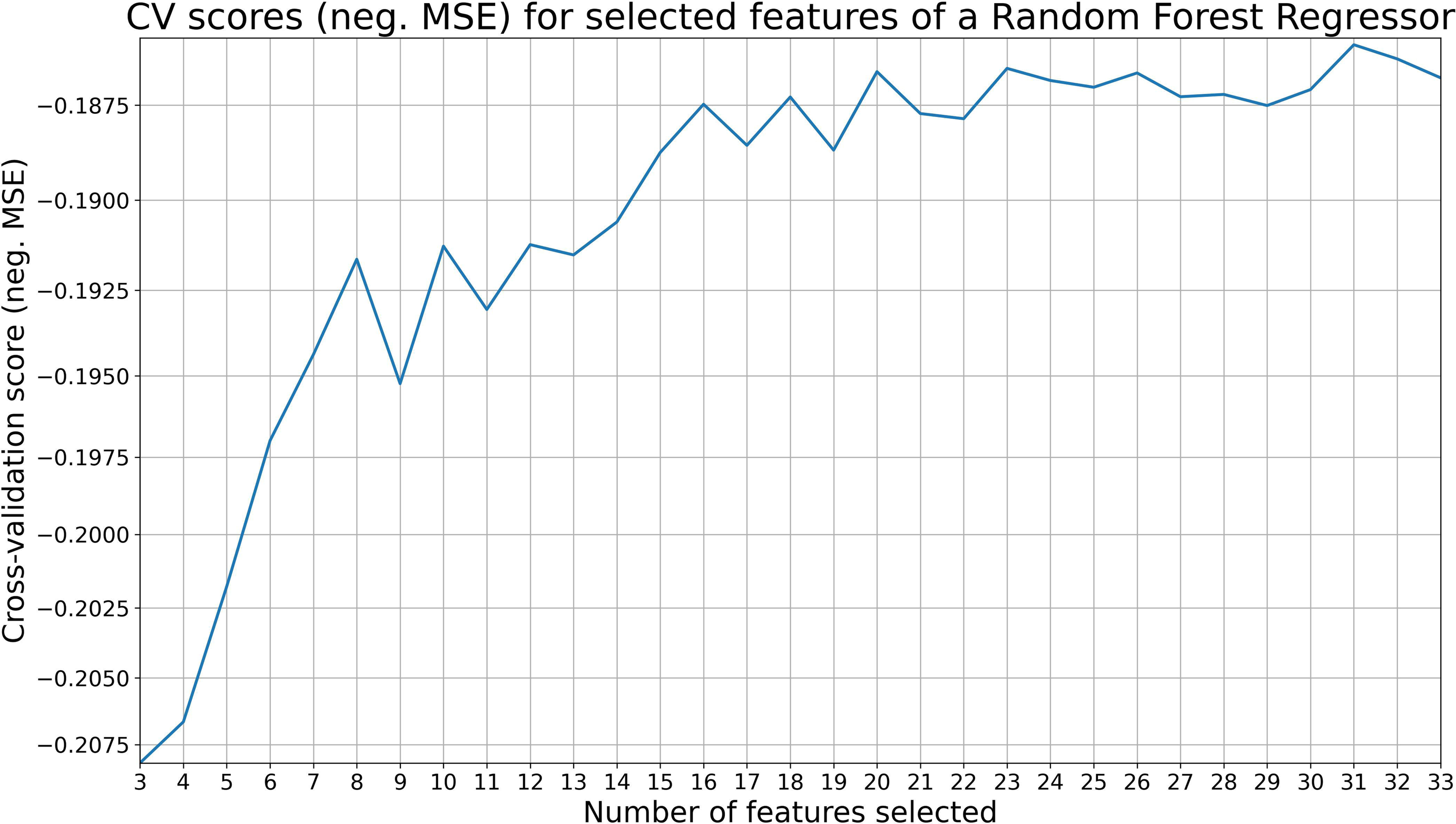
Cross-validation scores (negative MSE) for n number of features.

Feature ranking with recursive feature elimination with a random forest regressor was then used on the whole training set with the previously selected number of features to choose the most important features for regression. These features were used in all following machine learning analyses (hyperparameter tuning and fitting) described below.

### Tuning of Machine-Learning Algorithms and Model Fitting

We used a gradient boosting regressor (GBR), k-nearest neighbors regressor (KNN), random forest regressor (RFR), support vector regression (SVR), multi-layer perceptron regression (MLP) as machine learning algorithms, and multivariate linear regression (MLR) as reference. All features (independent variables) and labels (dependent variable) were standardized before training by removing the mean and scaling them to unit variance, using the formula 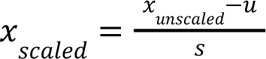, where *u* is the mean and *s* is the standard deviation of the training or test data (see Figure 2).

Hyperparameter tuning was performed within the training set using a 5-fold group cross-validation with a grid search approach, i.e., an exhaustive search over all (combinations of) prespecified parameter values. Negative MSE was again used as a performance metric. See S2 Appendix for the default parameters, the hyperparameter choices and tuned parameter values.

After hyperparameter tuning, each regressor was run again on the whole training set using the selected hyperparameter values and the selected features (see section ‘Feature Selection’); see Figure 2. For comparison, each regressor was also run with its default hyperparameter values.

For most accurate and unbiased feature importances, we used the drop column method and calculated the root mean squared error (RMSE) difference with 10 repetitions of a 5-fold group cross-validation for each feature [23].

### Performance Evaluation

Spearman’s rank correlation coefficient was calculated on averaged data per patient. Thus, we used this correlation measure between the patient’s averaged measured paO_2_ and the averaged parameters FiO_2_, pAO_2,_ and Gadrey’s paO_2_, respectively. These correlations were used as baseline values, i.e., as performances which can be simply achieved by assessing paO_2_ through single parameters, which should be significantly improved using machine learning.

For each algorithm, performance as measured by Spearman’s rank correlation between the predicted and the measured value was compared to the aforementioned correlations for both default and tuned hyperparameter values. Spearman’s ρ was calculated as 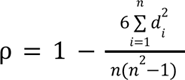 with *d_i_* being the difference between the two ranks of observation *i* (here: average per patient) and *n* being the number of (test) observations.

Additionally to Spearman’s rank correlation coefficient, performance evaluation was also conducted using the mean absolute percentage error (MAPE) and the RMSE, yielding a performance matrix with three columns. MAPE was calculated as 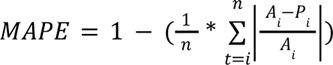 where *i* is the index of the (test) observation; *A_i_* stands for the actual value, and *P_i_* stands for the predicted value. RMSE was calculated as 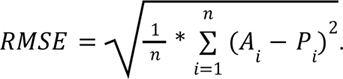

For each algorithm, we ranked the tuned estimator from 1 for best to 6 for worst based on the test set within each quality measure of the performance matrix (MAPE, Spearman’s ρ and RMSE). The best algorithm was determined by the lowest overall score, which was the sum of all ranks per estimator.

Outliers in these predictions were defined as their percentage error values 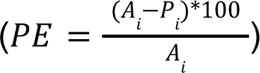 that were either smaller than three times of interquartile range (IQR) plus the first quartile (Q1) or larger than three times of IQR plus the third quartile (Q3). We investigated all corresponding observations to detect differences between highly over- or underestimated paO_2_ values.

### Implementation, Reproducibility, and Reporting

Data extraction was performed in Python 3.8.0, statistical analyses were performed in Python 3.9.5 on a macOS 10.16 machine. We used the following libraries for these tasks using a random state of 42 wherever applicable:

- Data preparation: pandas (version: 1.3.4)
- Data visualization: matplotlib (3.5.0), seaborn (0.11.2)
- Missing data imputations: scikit-learn (version: 1.0.1)
- Feature selection: pandas, numpy (1.21.4), scikit-learn
- Hyperparameter tuning: scikit-learn, numpy
- Statistical tests: numpy, scipy (1.7.2), statsmodels (0.13.2)

The code for the analysis in this paper is provided in the online appendix for the purpose of reproducibility. Several Jupyter Notebooks files (performed with IPython 7.29.0, jupyter_client 7.0.6, jupyter_core 4.9.1, notebook 6.4.6) are provided to fully reproduce the results of the paper. For more detailed instructions please refer to the readme file in the online appendix. Note that for the MLP learner results may differ slightly due to its parallel computing implementation.

Correlations were assessed using Spearman’s rank correlation coefficient on averaged values per patient. Differences between two groups were assessed using the Mann-Whitney U rank test. Significance threshold was set to the 5% level.

Extensive reporting for the prediction model development was done using the Transparent Reporting of a multivariable prediction model for Individual Prognosis Or Diagnosis (TRIPOD) checklist (see S3 Appendix). The Strengthening the Reporting of Observational Studies in Epidemiology (STROBE) guidelines were used to report results of the study (see S4 Appendix).

## RESULTS

### Patient Cohort

During the study period, 6,409 intracranial surgeries (number of surgeries, N) with 24,605 observations (number of perioperative value sets comprising one ABG and all corresponding ventilation and surgical parameters as well as demographic data and vital signs, n) met the inclusion criteria. We removed 71 observations from 16 surgical interventions due to invalid case numbers and thus, missing documentation in the hospital information system. An additional 28 interventions (217 observations) were excluded due to missing FiO_2_, CO_2_, initial paO_2_/FiO_2_ ratio (Horowitz index), SpO_2_, or length-of-stay values. Another 2,185 interventions were excluded based on our other criteria, resulting in a final data set of 4,180 surgeries with 12,497 observations (see Figure 1).

The mean age was 53.1 years and 56.3 % were female patients. The mean incision to suture time was 282 min, with a mean ventilation time of 406 min. The mean initial Horowitz index was 464.1 mmHg, the mean hemoglobin value was 12.1 g/dl and the mean body mass index was 25.2 kg/m^2^.

### Missing Data

Most features had complete and high quality data. Only preoperative creatinine and, to a much lower extent, ventilation parameters were affected. We iteratively imputed the missing values of respiratory rate (seven values), compliance (seven values), respiratory minute volume (seven values), blood pressure and heart rate (four values each), and blood pH values (two values), hemoglobin (twelve values), and creatinine (1,659 values).

### Feature Selection

The calculated negative MSE during cross-validated RFE for the scaled training set is shown in Figure 2. The calculated optimal number of features was 31 (neg. MSE = -0.1860 mmHg^2^). The second highest cross-validated scores were reached with 20 and 23 features. Thus, we selected 20 features (neg. MSE = -0.1866 mmHg^2^) to be used with all algorithms (see Figure 3). The descriptive statistics of the 20 selected features are shown in Table 1.

**Table 1.**
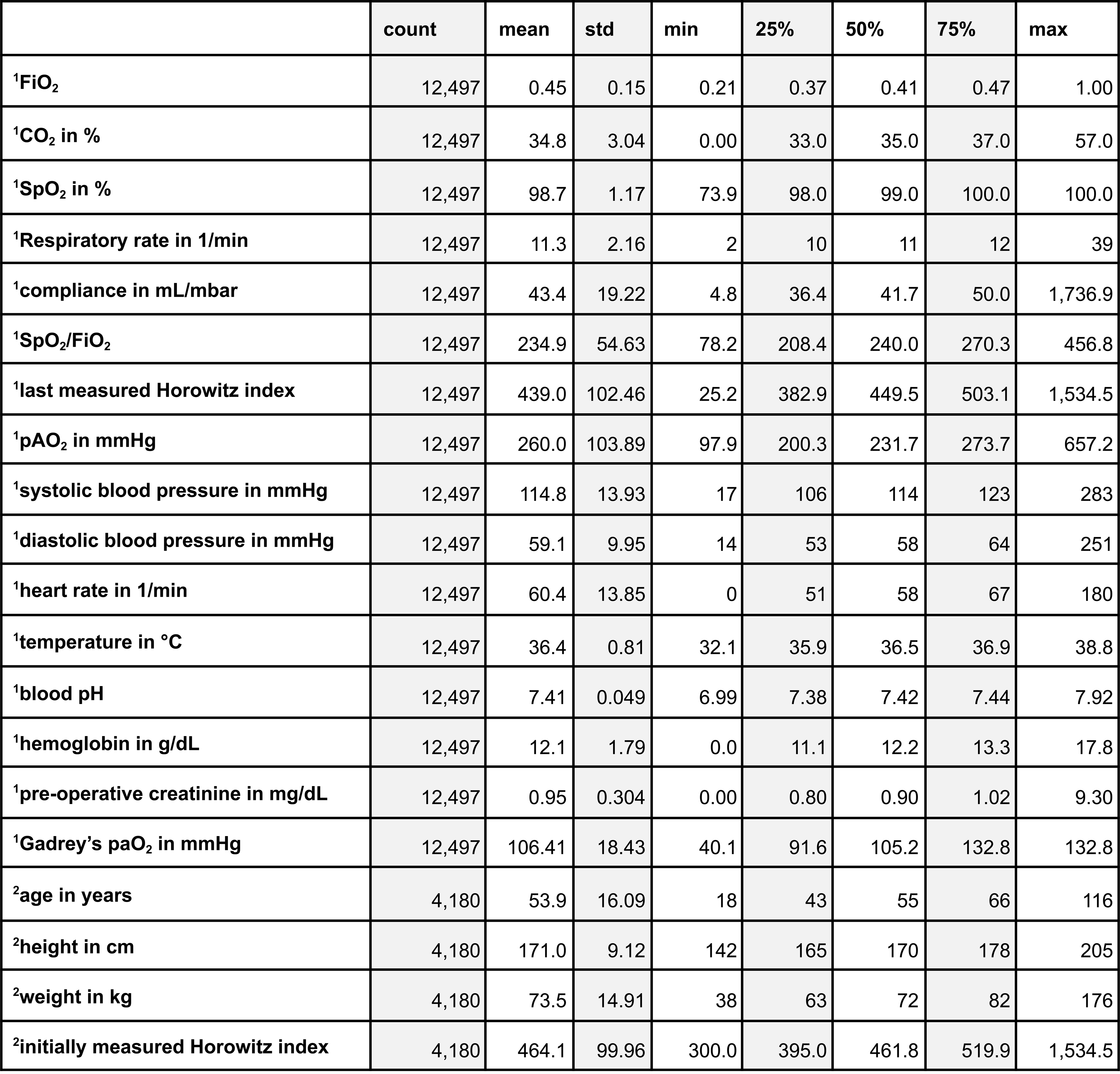
Description of selected features for paO_2_ prediction ^1^ per feature (if multiple measurements per patients) or ^2^ per patient (if one-time measurement)

|  | count | mean | std | min | 25% | 50% | 75% | max |
| --- | --- | --- | --- | --- | --- | --- | --- | --- |
| <sup>1</sup>FiO<sub>2</sub> | 12,497 | 0.45 | 0.15 | 0.21 | 0.37 | 0.41 | 0.47 | 1.00 |
| <sup>1</sup>CO<sub>2</sub> in % | 12,497 | 34.8 | 3.04 | 0.00 | 33.0 | 35.0 | 37.0 | 57.0 |
| <sup>1</sup>SpO<sub>2</sub> in % | 12,497 | 98.7 | 1.17 | 73.9 | 98.0 | 99.0 | 100.0 | 100.0 |
| <sup>1</sup>Respiratory rate in 1/min | 12,497 | 11.3 | 2.16 | 2 | 10 | 11 | 12 | 39 |
| <sup>1</sup>compliance in mL/mbar | 12,497 | 43.4 | 19.22 | 4.8 | 36.4 | 41.7 | 50.0 | 1,736.9 |
| <sup>1</sup>SpO<sub>2</sub>/FiO<sub>2</sub> | 12,497 | 234.9 | 54.63 | 78.2 | 208.4 | 240.0 | 270.3 | 456.8 |
| <sup>1</sup>last measured Horowitz index | 12,497 | 439.0 | 102.46 | 25.2 | 382.9 | 449.5 | 503.1 | 1,534.5 |
| <sup>1</sup>pAO<sub>2</sub> in mmHg | 12,497 | 260.0 | 103.89 | 97.9 | 200.3 | 231.7 | 273.7 | 657.2 |
| <sup>1</sup>systolic blood pressure in mmHg | 12,497 | 114.8 | 13.93 | 17 | 106 | 114 | 123 | 283 |
| <sup>1</sup>diastolic blood pressure in mmHg | 12,497 | 59.1 | 9.95 | 14 | 53 | 58 | 64 | 251 |
| <sup>1</sup>heart rate in 1/min | 12,497 | 60.4 | 13.85 | 0 | 51 | 58 | 67 | 180 |
| <sup>1</sup>temperature in °C | 12,497 | 36.4 | 0.81 | 32.1 | 35.9 | 36.5 | 36.9 | 38.8 |
| <sup>1</sup>blood pH | 12,497 | 7.41 | 0.049 | 6.99 | 7.38 | 7.42 | 7.44 | 7.92 |
| <sup>1</sup>hemoglobin in g/dL | 12,497 | 12.1 | 1.79 | 0.0 | 11.1 | 12.2 | 13.3 | 17.8 |
| <sup>1</sup>pre-operative creatinine in mg/dL | 12,497 | 0.95 | 0.304 | 0.00 | 0.80 | 0.90 | 1.02 | 9.30 |
| <sup>1</sup>Gadrey's paO<sub>2</sub> in mmHg | 12,497 | 106.41 | 18.43 | 40.1 | 91.6 | 105.2 | 132.8 | 132.8 |
| <sup>2</sup>age in years | 4,180 | 53.9 | 16.09 | 18 | 43 | 55 | 66 | 116 |
| <sup>2</sup>height in cm | 4,180 | 171.0 | 9.12 | 142 | 165 | 170 | 178 | 205 |
| <sup>2</sup>weight in kg | 4,180 | 73.5 | 14.91 | 38 | 63 | 72 | 82 | 176 |
| <sup>2</sup>initially measured Horowitz index | 4,180 | 464.1 | 99.96 | 300.0 | 395.0 | 461.8 | 519.9 | 1,534.5 |

The feature ranking with recursive feature elimination based on 20 features returned FiO_2_, CO_2_, SpO_2_, respiratory rate, compliance, first perioperative paO_2_/FiO_2_ ratio (Horowitz index), the last measured Horowitz index, alveolar oxygen partial pressure (pAO_2_, calculated by the alveolar gas equation), systolic blood pressure, diastolic blood pressure, heart rate, temperature, blood pH, hemoglobin value, preoperative creatinine, age, height, weight, SpO_2_/FiO_2_ ratio, and Gadrey’s paO_2_.

### Comparison to Popular Proxies

The correlations of averaged FiO_2_, the pAO_2_, and Gadrey’s paO_2_ to the averaged measured paO_2_ were ρ=0.6781 (95 % confidence interval (CI): [0.6703; 0.6858]), ρ=0.6779 (95 % CI: [0.6701; 0.6856]) and ρ=0.2850 (95 % CI: [0.2718; 0.2981]). All base regressors for paO_2_ prediction (not-tuned regressors with default parameters) reached MAPEs between 10 and 13 %, RMSEs of less than 38 mmHg (for rescaled data) and correlation coefficients between 0.8207 and 0.8988 with confidence intervals ranging from 0.8100 to 0.90471. Thus, all base regressors showed a significantly better ability to approximate measured paO_2_ values than Gadrey’s paO_2_, FiO_2_ settings, or pAO_2_ alone.

### Tuned Algorithms

After hyperparameter tuning using a cross-validated grid search approach with neg. MSE as performance metric (see Appendix S2), the gradient boosting regressor (GBR) had an increased learning rate (0.1325) and decreased number of boosting stages (65). The minimum number of samples required to split an internal node was set to nine, while the minimum number of samples required to be a leaf node was set to four. The Huber function was used as the loss function.

The tuned k-nearest neighbors regressor (KNN) used 20 neighbors and a BallTree algorithm to determine these neighbors. The used leaf size was cut in half to 15. The Manhattan distance was used for the power parameter in the Minkowsi metric.

The final random forest regressor (RFR) increased the number of trees to 150 and the minimum number of samples needed for a leaf node to 8. The maximum depth of the tree was set to 23.

Only three parameters changed in the tuned support vector regression (SVR). The regularization parameter, C, was increased to 1.76 while epsilon was decreased to 0.01. Neither the kernel function nor the kernel coefficient were changed.

In the multi-layer perceptron regressor (MLP), the hyperbolic tan function was used for activation. The L2 penalty parameter alpha was increased to 1, and the solver for weight optimization was set to a method based on a stochastic gradient-based optimizer [24]. We used one hidden layer with 24 hidden neurons.

### Mean Absolute Percentage Error, Spearman’s Correlation Coefficient, and Mean Squared Error

The KNN reached the worst MAPE with 13.23 %, followed by the MLR with 11.69 %. The SVR performed a little bit better (11.77 %). Less than 11% were reached by the MLP (10.50 %). The GBR (10.45 %) and the RFR (10.35 %). For this performance metric, the RFR yielded the best result (see Table 1).

The lowest Spearman’s correlation coefficient was found for the predictions made with the KNN (ρ=0.8604). Better performed the linear regression, the SVR, and the MLP with ρ=0.8664, ρ=0.8737, and ρ=0.8965, respectively. Both the RFR and GBR reached correlations over 0.90 (0.9017 and 0.9029). For this performance metric, the GBR yielded the best result (see Table 1).

The highest RMSE by far was calculated for the KNN (RMSE=34.22 mmHg). A slightly lower value was found for the SVR with RMSE=33.65 mmHg and the linear regression with RMSE=33.56 mmHg. All RMSEs of MLP, RFR, and GBR were very close to each other (MLP: 30.69 mmHg, RFR: 30.86 mmHg, GBR=31.31 mmHg). For this performance metric, the MLP yielded the best result (see Table 1).

In summary, RFR performed best by reaching the lowest sum of ranks in our performance matrix from Table 2 (5), but was closely followed by GBR (6) and MLP (7).

**Table 2.**
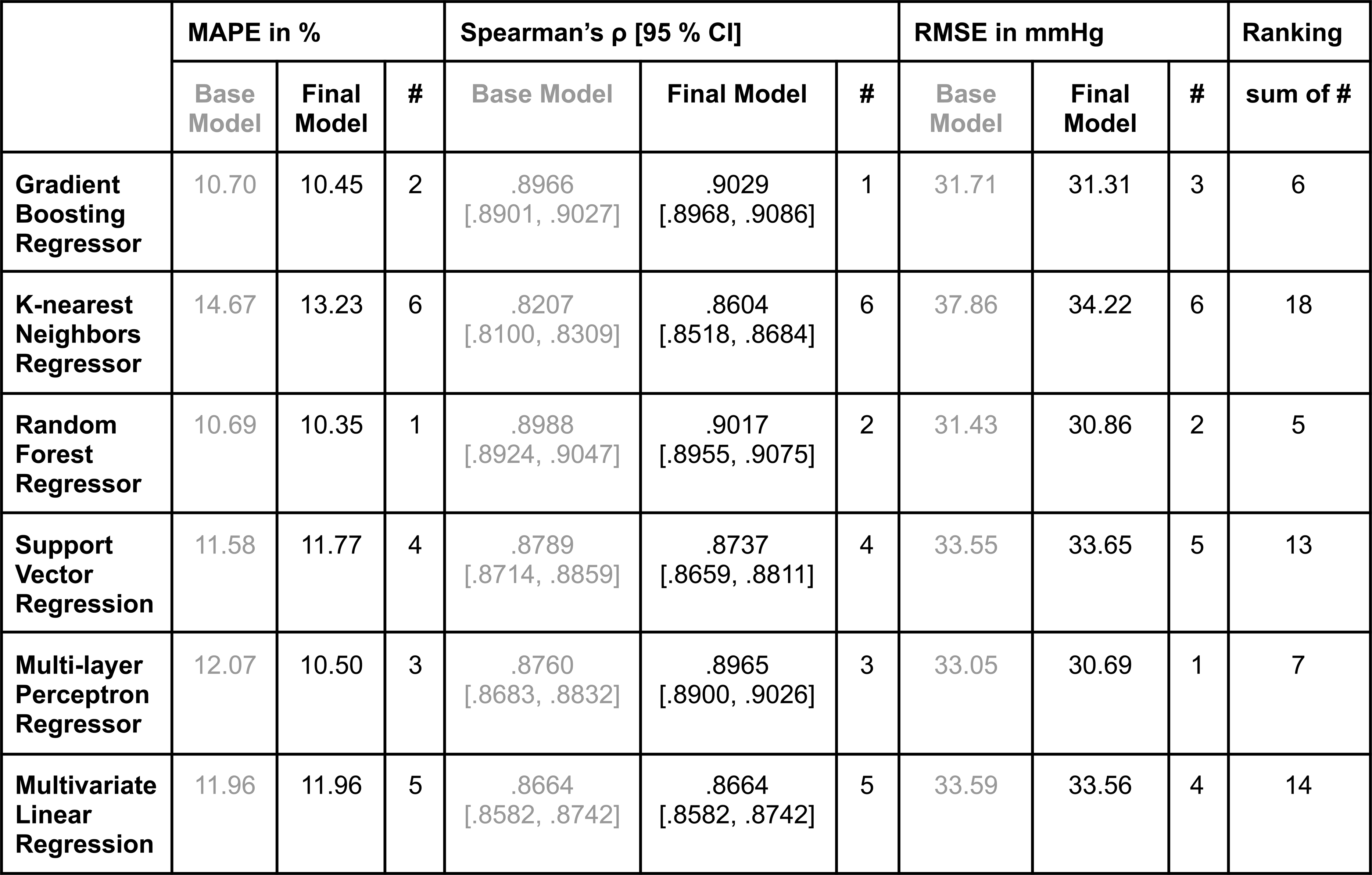
MAPE, Spearman’s ρ [95 % CI], and RMSE for each algorithm with default parameter values (“Base model”) and tuned parameter values (“Final model”) based on the test data set. The rank of the regressor for the considered performance metric is indicated in the column “#”.

To visualize their correlation for each algorithm, the predicted and measured paO_2_ values of the test set were plotted against each other (see Figure 4).

**Figure 4.**
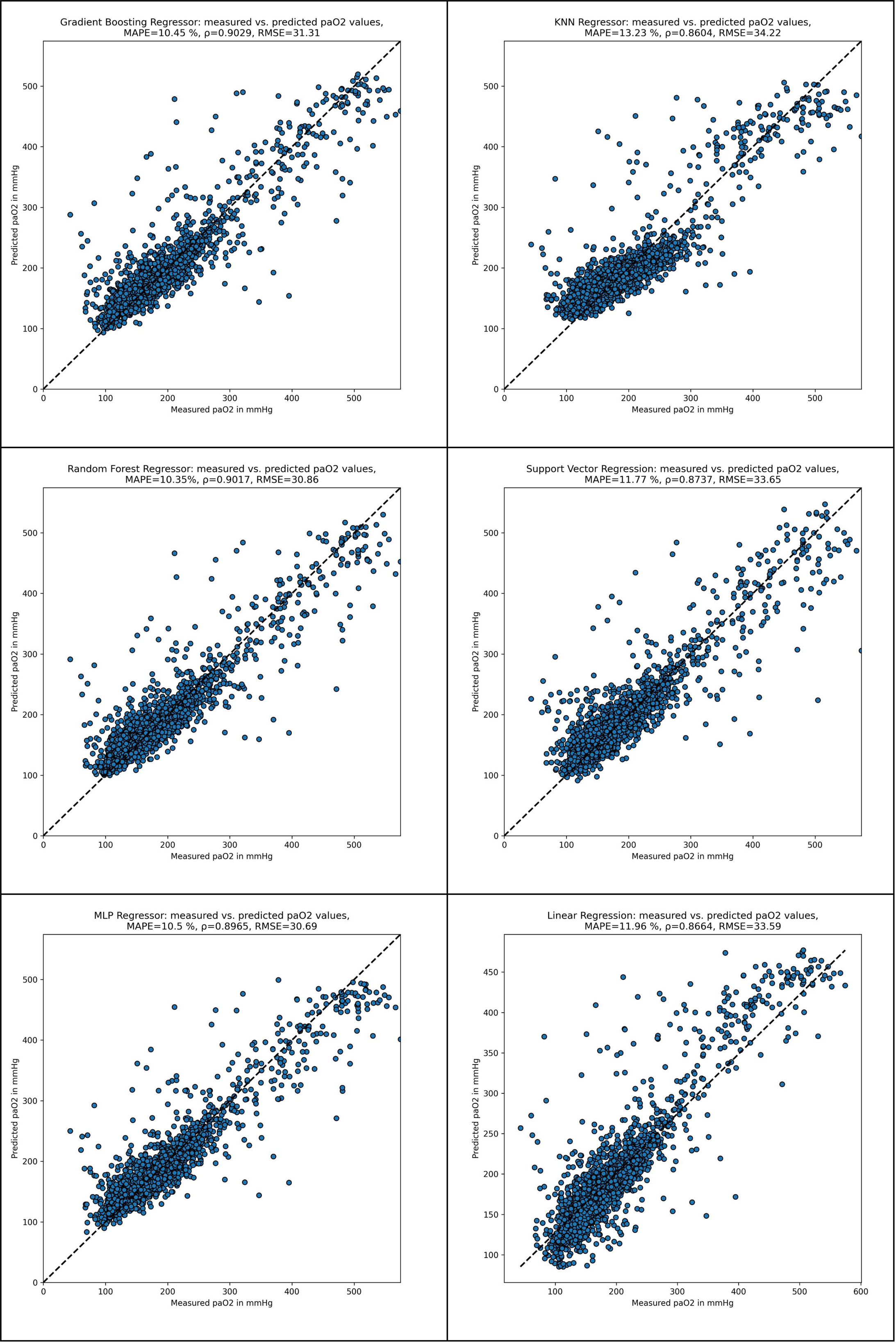
Measured vs. predicted paO_2_ values for different estimators. A. Gradient Boosting Regressor, B. k-nearest Neighbors Regressor, C. Random Forest Regressor, D. Support Vector Regression, E. Multi-layer Perceptron Regressor, F. Multivariate Linear Regression

We calculated a cross-validated feature importance with 10 repetitions for the RFR, which is shown in Figure 5.A. The most important feature was the last measured Horowitz index (RMSE difference [diff_RMSE_]=1.14 mmHg), followed by age (diff_RMSE_=0.32 mmHg). Less important were weight (diff_RMSE_=0.12 mmHg), preoperative creatinine (diff_RMSE_=0.12 mmHg), and the initially measured Horowitz index (diff_RMSE_=0.10 mmHg). All other features had lower RMSE differences, SpO_2_, height, systolic blood pressure, hemoglobin value, respiratory rate, FiO_2,_ and SpO_2_/FiO_2_ yielded negative RMSE differences (see Figure 5.A).

We investigated all features for the predicted paO_2_ values that were more than three times of the interquartile range (IQR) beyond the first (Q1) and third (Q3) quartiles of the overall percentage errors (Q1-3*IQR: -40.0 %, Q3+3*IQR: 39.8 %, see Figure 5.B). Among these outliers (see Figure 5.C), significant differences (p<0.05) in their distribution were found in eight features.

**Figure 5.**
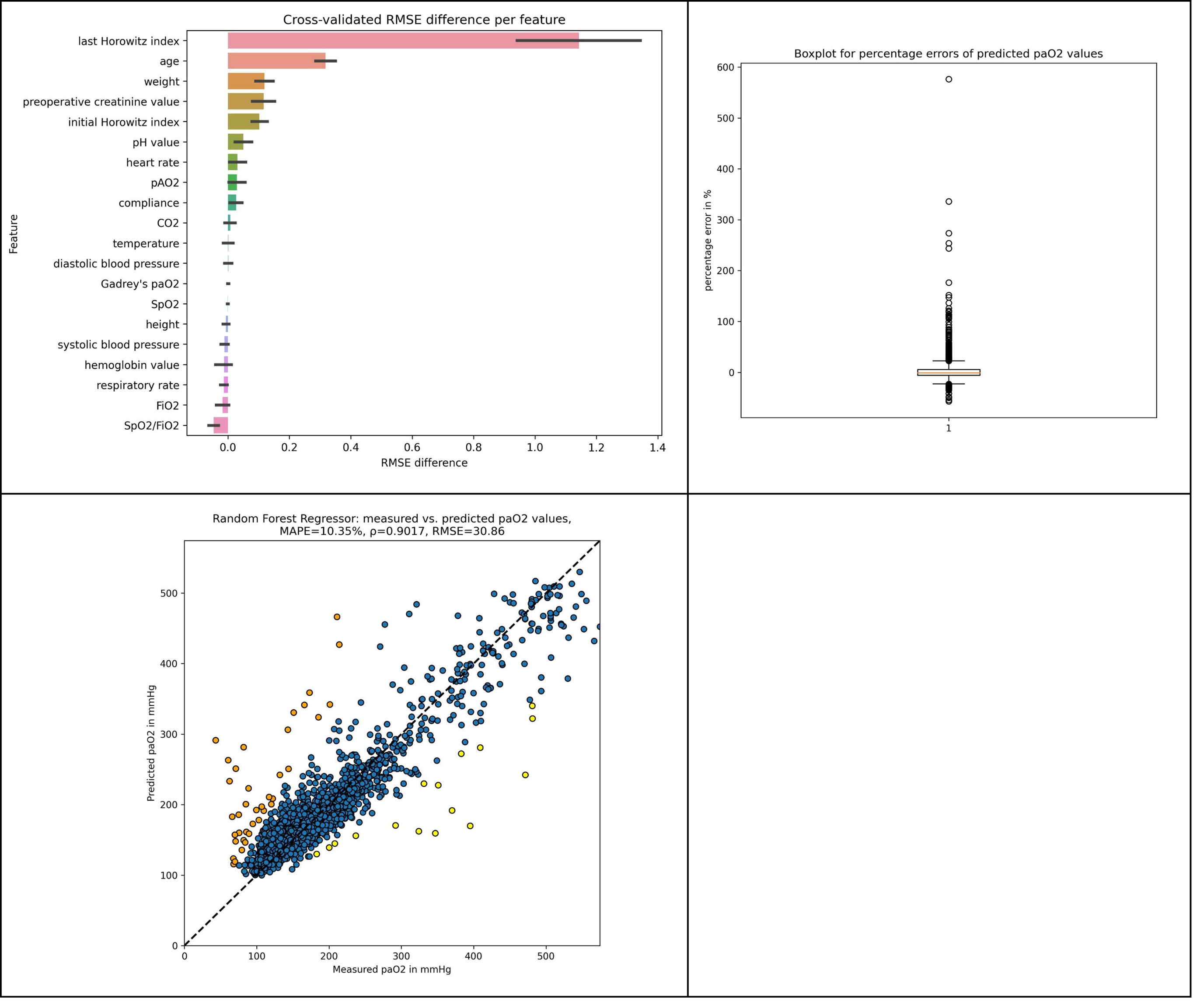
Random Forest Regressor. A. Feature importance by cross-validated MSE difference with 95 % CI, B. Boxplot of percentage errors for predicted paO_2_ values, C. Scatter Plot of measured versus predicted paO_2_ values. Orange: predicted paO_2_ values whose percentage error was larger than the 75th percentile plus three times of interquartile range of the overall percentage error (n=38). Yellow: predicted paO_2_ values whose percentage error was lower than the 25th percentile minus three times of interquartile range of the overall percentage error (n=16).

The median CO_2_ value in the overestimated group was 34.8 % compared to 34.3 % in the underestimated group (p=0.0137). Median values of compliance (43.4 ml/mbar vs. 41.7 ml/mbar, p=0.0089), the initially measured Horowitz index (464.5 vs. 438.8, p<0.0001) and the last measured Horowitz index (434.7 vs. 410.7, p<0.0001) were all greater in the overestimated group, whereas the medians of respiratory rate (11.1/min vs. 11.5/min, p=0.0132), age (52.5 vs. 56.6, p<0.0001), heart rate (59.6/min vs. 62.4/min, p=0.0001) and weight (72.3 kg vs. 76.6 kg, p<0.0001) were all greater in the underestimated group.

## DISCUSSION

Assessment of paO_2_ on a semi-continuous basis is currently impossible. Frequent BGAs are time-costly and potentially harmful for the patient due to risk of infection and blood loss. However, paO_2_ is one of the few tools available to estimate a patient’s degree of hyperoxygenation. In this manuscript, we therefore successfully trained, tuned, and compared several machine learning algorithms to estimate continuous paO_2_ values in neurosurgical patients. The random forest regressor performed best over all three categories of the performance matrix and thus, was able to predict intraoperative paO_2_ values based on 20 features with 10.35 % of mean absolute error, a correlation of 0.9017 and a root mean squared error of 30.86 mmHg. Still, both the gradient boosting regressor as well as the multi-layer perceptron regressor performed minimally worse and scored second and third. With our work, it is possible to constantly draw conclusions about tissue oxygenation by calculating a synthetic paO_2_ value without the need of an additional ABG. In addition to the non-invasive monitoring of hypoxemia, anaesthesiologists are now enabled to identify hyperoxemia at an early stage and thus, take precautions to avoid excessive oxygenation.

To the best of our knowledge, this is the first study to successfully predict live paO_2_ values. In the past, FiO_2_ or the alveolar oxygen partial pressure have been used to extrapolate to a corresponding paO_2_ value. In our data, the correlation between these and the measured paO_2_ values was less than 0.65, which was significantly worse than all our machine learning algorithms. Gadrey et al. were the first to introduce a new equation of paO_2_ calculation, based on two constant values and the SpO_2_ value [17]. With SpO_2_ being the only measured variable, the outcome has a natural maximum of 132.8 mmHg. Thus, the equation is not suitable to model hyperoxia. Additionally, the correlation coefficient was at 0.31, which was even worse than FiO_2_ or the pAO_2_, despite the fact it was the fifth most important feature for RFR.

Neurosurgical patients are particularly well suited to test different algorithms for the prediction of paO_2_ during surgery for different reasons: First, there is usually an isolated pathology in the neurocranium, and no surgical procedure affects the thorax and therefore gas exchange remains unaffected. Assuming a normal lung function, a conclusion can be drawn from blood to tissue oxygenation. Second, arterial catheters are part of the standard monitoring procedures for patients undergoing craniotomy. They are mainly used for continuous blood pressure monitoring, but consecutively also for ABGs. Third, due to safety precautions, there might be a tendency to use a higher FiO_2_ than needed to ensure tissue oxygenation in the brain as the patient’s airway during many neurosurgical interventions is not accessible. Consequently, these patients are a suitable focus group for close monitoring of blood oxygenation through synthetic paO_2_ values to avoid not only hypoxia but also hyperoxia. Additionally, as craniotomies usually take three to five hours of surgery time, several ABGs are taken in the meantime providing a profound data lake.

Our machine learning algorithms can model even complex interactions between perioperative and sociodemographic values. It is not surprising that the Random Forest Regressor performed best. Random forests often perform well with big data (large n) and high dimensionality (many variables), they are robust to outliers and non-linear data and can handle unbalanced data. In addition, they tend to have a low bias with a moderate variance. Their only drawbacks lay in the need for a lot of memory and the lack of interpretability [25].

The most important feature (Figure 5.A) was the last measured Horowitz index. This comes as no surprise as the last measured paO_2_ value is the numerator in this index. Interestingly, age is the second most important feature in the RFR. PaO_2_ is known to decrease with age; this is accompanied by several changes in the mechanical properties of the lung, e.g., loss of elastic recoil and increase in closing volume [26, 27]. Additionally, it is known that in the elderly, aging increases sensitivity to oxidative stress and thus shifts the demand of O_2_ and related toxicity [28].

Some studies found that heavier patients have lower paO_2_ values, which explains weight being the third most important feature (and second non-ventilation setting related one) [27, 29].

Prnjavorac et al. analyzed the significance of serum creatinine levels in respiratory failure, noticing that lower creatinine levels were associated with improvements of respiratory status [30]. Their findings might explain why the preoperative creatinine value was the fourth most important feature.

Still, Decision Trees (as part of Random Forests) and (more complex) Machine Learning algorithms in general are not known for their interpretability per se, although they can reach high accuracies [31].

Both recursive feature elimination algorithms used the implemented feature importance function of the Random Forest Regressor in the sklearn Python library. As this feature importance is impurity-based, it can be misleading for high cardinality features [32]. Thus, we tried to remove this bias by calculating a drop-column-based feature importance. As this was a different approach, the resulting feature importance could show negative RSMEs as shown in Figure 5.

This study faces some limitations. First, all of our patients had an initially measured Horowitz index of more than 300, which was chosen to avoid including patients with relevant lung injury [33]. Thus, our findings may not be generalizable to patients with relevant pulmonary dysfunction, such as chronic obstructive pulmonary disease, asthma, lung cancer, or even acute respiratory distress syndrome. Second, besides the surgical procedure and the main diagnosis, we did not take any other comorbidities into account when training and fitting the different algorithms. The reason for this was the need to develop a generalizable algorithm, which could be used in a broad range of patients with different kinds of comorbidities but the same type of surgical procedure. Still, with 10 % of MAPE we yielded very accurate results. Additionally, we only included patients with mechanical ventilation and general anesthesia who received at least two ABGs. Thus, our results may not be applicable for patients with small interventions that require local anesthesia only, meaning also no intubation is needed and no ABG is taken.

Additionally, in the range of normoxic paO_2_ values (60-120 mmHg), we rather overestimate the paO_2_ value, whereas in severe hyperoxia (more than 300 mmHg) we rather underestimate the true value (see Figure 4C).

Another limitation were the selected features in the recursive feature elimination process. We selected the random forest regressor as an estimator, as the RFE algorithm required an estimator with an implemented feature importance property and not all algorithms provided this functionality. Thus, the features selected by a random forest regressor were used in each of the six algorithms. Still, as tree-based algorithms naturally compute the decrease of impurity with each feature, the selected features are rather generalizable.

Our research shows that machine learning algorithms can be utilized to precisely predict paO_2_ values for a range between 100 to 300 mmHg. Our RFR did not only achieve the lowest MAPE of around 10 %, but also the second highest correlation coefficient and the second smallest RMSE. Although some papers exist that extrapolate paO_2_ values, this is the first model for online prediction of paO_2_ values with satisfactory results. Besides the non-invasive measurement of SpO_2_ to monitor hypoxemia, anaesthesiologists are now able to monitor hyperoxemia as well, without the need of additional ABGs.

Future studies are needed to validate our method in other patient collectives like neurosurgical patients with acute respiratory distress syndrome as well as for prospective validation in the same patient collective. Neurosurgical patients are of particular interest, as reperfusion and ischemia are two vital challenges. Additionally, most of the studies on vasoconstrictors were performed in either healthy volunteers or under stable conditions [34]. Thus, these patients can be used to study the effect of hyperoxia, based on calculated paO_2_ values.

## Data Availability

All data produced in the present study are available upon reasonable request to the authors.

## ABBREVIATIONS

ABG: Arterial Blood Gas
CI: Confidence Interval
diff_MSE_: MSE Difference
GBR: Gradient Boosting Regressor
IQR: Interquartile Range
KNN Regressor: K-nearest Neighbors Regressor
MLR: Multivariate Linear Regression
MAPE: Mean Absolute Percentage Error
MLP Regressor: Multi-layer Perceptron Regressor
MLR: Multivariate Linear Regression
MSE: Mean Squared Error
n: Number of Observations (multiple per surgery)
N: Number of Surgeries
neg.: Negative
paO_2_: Partial Arterial Pressure of Oxygen
pAO_2_: Alveolar Oxygen Partial Pressure
Q1/Q3: First/Third Quartile
RFR: Random Forest Regressor
RMSE: Root Mean Squared Error
SVR: Support Vector Regression

## ACKNOWLEDGMENTS

We would like to thank Anna Jacob for proofreading.

## APPENDIX

### S1: Full List of Features

- pao2_measured: measured paO_2_ value in mmHg (continuous value),
- idx: incrementing number of the measurement (discrete value),
- fio2: inspiratory fraction of oxygen (continuous value),
- co2: end-tidal carbon dioxide in mmHg (continuous value),
- spo2: peripheral capillary oxygen saturation in % (continuous value),
- amv: respiratory minute volume in liter (continuous value),
- af: respiratory rate in 1/minute (discrete value),
- compliance: ventilation compliance in milliliter/millibar (continuous value),
- last_horowitz: last measured paO_2_/FiO_2_ ratio in mmHg (continuous value),
- horowitz_0: first paO_2_/FiO_2_ ratio in mmHg (continuous value),
- pao2_calc: pAO2 calculated value by the alveolar gas equation in mmHg (continuous value),
- systole: systolic value in mmHg (discrete value),
- diastole: diastolic value in mmHg (discrete value),
- mean_art_press: mean arterial pressure in mmHg (discrete value),
- pulse: heart rate in 1/minute (discrete value),
- temp: temperature value in degree celsius (continuous value),
- ph: last measured pH-value in (continuous value),
- hb: last measured hemoglobin value in (continuous value),
- krea_0: preoperative creatinine value in milligram/deciliter (continuous value),
- age: age in years (continuous value),
- height: height in centimeters (discrete value),
- weight: weight in kilogram (value),
- already_intubated: indicates whether a patient was already intubated before surgery (dichotomous value),
- time_to_cut: time from intubation to cut in minutes (continuous value),
- spo2_fio2: spO_2_/FiO_2_ ratio (continuous value),
- gadrey: paO_2_ values calculated by Gadrey et al. in mmHg[17](continuous value),
- sex_m: indicates a male patient (dichotomous value),
- sex_w: indicates a female patient (dichotomous value),
- asa_1: indicates ASA score I (dichotomous value),
- asa_2: indicates ASA score II (dichotomous value),
- asa_3: indicates ASA score III (dichotomous value),
- asa_4: indicates ASA score IV (dichotomous value),
- asa_5: indicates ASA score V (dichotomous value),
- timepoint_of_measurement_intraoperative: indicates whether the measurement was done intraoperatively (dichotomous value),
- timepoint_of_measurement_post-surgery: indicates whether the measurement was done postoperatively (dichotomous value),
- timepoint_of_measurement_pre-surgery: indicates whether the measurement was done preoperatively (dichotomous value)

### S2: Default and Final Parameters of Estimators

#### Gradient Boosting Regressor

Default parameters:

- loss=’squared_error’,
- learning_rate=0.1,
- n_estimators=100,
- subsample=1.0,
- criterion=’friedman_mse’,
- min_samples_split=2,
- min_samples_leaf=1,
- min_weight_fraction_leaf=0.0,
- max_depth=3,
- min_impurity_decrease=0.0,
- min_impurity_split=None,
- init=None, random_state=None,
- max_features=None,
- alpha=0.9,
- verbose=0,
- max_leaf_nodes=None,
- warm_start=False,
- validation_fraction=0.1,
- n_iter_no_change=None,
- tol=0.0001,
- ccp_alpha=0.0

Hyperparameters:

- loss: [’squared_error’, ’huber’],
- learning_rate: array([0.01 , 0.1325, 0.255 , 0.3775, 0.5]),
- n_estimators: [50, 65, 80, 95],
- subsample: array([0.1 , 0.55, 1.]),
- criterion: [’friedman_mse’, ’squared_error’],
- min_samples_split: [3, 6, 9],
- min_samples_leaf: [2, 3, 4],
- max_features: [’auto’, ’sqrt’, ’log2’]

Changed parameters in best estimator:

- learning_rate: 0.1325,
- loss: ‘huber’,
- max_features: auto (here: 20),
- min_samples_leaf: 4,
- min_samples_split: 9,
- n_estimators: 65,

#### K-nearest Neighbors Regression

Default parameters:

- n_neighbors=5,
- weights=’uniform’,
- algorithm=’auto’,
- leaf_size=30,
- p=2,
- metric=’minkowski’,
- metric_params=None,
- n_jobs=None

Hyperparameters:

- n_neighbors: array([ 5, 10, 15, 20, 25, 30, 35, 40, 45, 50]),
- weights: [’uniform’],
- algorithm: [’ball_tree’, ’kd_tree’, ’brute’],
- leaf_size: array([15, 20, 25, 30, 35]),
- p: [1, 2]

Changed parameters in best estimator:

- algorithm: ’ball_tree’,
- n_neighbors: 20,
- leaf_size: 15,
- p: 1

#### Random Forest Regression

Default parameters:

- n_estimators=100,
- criterion=’squared_error’,
- max_depth=None,
- min_samples_split=2,
- min_samples_leaf=1,
- min_weight_fraction_leaf=0.0,
- max_features=’auto’,
- max_leaf_nodes=None,
- min_impurity_decrease=0.0,
- min_impurity_split=None,
- bootstrap=True,
- oob_score=False,
- n_jobs=None,
- random_state=42,
- verbose=0,
- warm_start=False,
- ccp_alpha=0.0,
- max_samples=None

Hyperparameters:

- n_estimators: [50, 66, 83, 100, 116, 133, 150, 166, 183, 200],
- max_features: [’auto’, ’sqrt’, ’log2’],
- max_depth: [2, 9, 16, 23, 30, None],
- min_samples_leaf: array([2, 5, 8]),
- bootstrap: [True]

Changed parameters in best estimator:

- max_depth: 23,
- min_samples_leaf: 8,
- n_estimators: 150

#### Support Vector Regression

Default parameters:

- kernel=’rbf’,
- degree=3,
- gamma=’scale’,
- coef0=0.0,
- tol=0.001,
- C=1.0,
- epsilon=0.1,
- shrinking=True,
- cache_size=200,
- verbose=False,
- max_iter=- 1

Hyperparameters:

- C: array([ 0.1, 1.75555556, 3.41111111, 5.06666667, 6.72222222, 8.37777778, 10.03333333, 11.68888889, 13.34444444, 15.]),
- epsilon: array([1.00000000e-02, 1.67555556e+00, 3.34111111e+00, 5.00666667e+00, 6.67222222e+00, 8.33777778e+00, 1.00033333e+01, 1.16688889e+01, 1.33344444e+01, 1.50000000e+01]),
- gamma: [’auto’, ’scale’],
- kernel: [’linear’, ’poly’, ’rbf’, ’sigmoid’]

Changed parameters in best estimator:

- C: 1.7555555555555558,
- epsilon: 0.01,

#### Multi-layer Perceptron Regressor

- hidden_layer_sizes=100,
- activation=’relu’,
- solver=’adam’,
- alpha=0.0001,
- batch_size=’auto’,
- learning_rate=’constant’,
- learning_rate_init=0.001,
- power_t=0.5,
- max_iter=200,
- shuffle=True,
- random_state=None,
- tol=0.0001,
- verbose=False,
- warm_start=False,
- momentum=0.9,
- nesterovs_momentum=True,
- early_stopping=False,
- validation_fraction=0.1,
- beta_1=0.9,
- beta_2=0.999,
- epsilon=1e-08,
- n_iter_no_change=10,
- max_fun=15000 Hyperparameters: (39,), (40,), (41,), (45,), (50,), (56,), (64,), (75,), (90,), (113,), (151,), (227,), (454,)],
- activation: [’relu’, ’tanh’, ’logistic’, ’identity’],
- alpha: [1e-05, 0.0001, 0.001, 0.01, 0.1, 1, 10],
- learning_rate: [’constant’, ’adaptive’, ’invscaling’],
- solver: [’adam’, ’sgd’]

Changed parameters in best estimator:

- activation: tanh,
- alpha: 1,
- hidden_layer_sizes: (24,),
- solver: adam

#### Linear Regression

Default parameters:

- fit_intercept=True,
- normalize=False,
- copy_X=True,
- n_jobs=None,
- positive=False

Hyperparameters:

- fit_interceptss: [True],
- normalize: [False, True],
- copy_X: [False, True],
- positive: [False, True],

Changed parameters in best estimator: none

### S3: TRIPOD checklist

#### TRIPOD Checklist for Prediction Model Development

| Section/Topic | Item | Checklist Item | Page |
| --- | --- | --- | --- |
| **Title and abstract** |  |  |  |
| Title | 1 | Identify the study as developing and/or validating a multivariable prediction model, the target population, and the outcome to be predicted. | 1 |
| Abstract | 2 | Provide a summary of objectives, study design, setting, participants, sample size, predictors, outcome, statistical analysis, results, and conclusions. | 2 |
| **Introduction** |  |  |  |
| Background and objectives | 3a | Explain the medical context (including whether diagnostic or prognostic) and rationale for developing or validating the multivariable prediction model, including references to existing models. | 4 |
|  | 3b | Specify the objectives, including whether the study describes the development or validation of the model or both. | 5 |
| **Methods** |  |  |  |
| Source of data | 4a | Describe the study design or source of data (e.g., randomized trial, cohort, or registry data), separately for the development and validation data sets, if applicable. | 6-7 |
|  | 4b | Specify the key study dates, including start of accrual; end of accrual; and, if applicable, end of follow-up. | 6 |
| Participants | 5a | Specify key elements of the study setting (e.g., primary care, secondary care, general population) including number and location of centres. | 6 |
|  | 5b | Describe eligibility criteria for participants. | 6, 7 |
|  | 5c | Give details of treatments received, if relevant. | n.a. |
| Outcome | 6a | Clearly define the outcome that is predicted by the prediction model, including how and when assessed. | 5, 9-10 |
|  | 6b | Report any actions to blind assessment of the outcome to be predicted. | n.a. |
| Predictors | 7a | Clearly define all predictors used in developing or validating the multivariable prediction model, including how and when they were measured. | 13 |
|  | 7b | Report any actions to blind assessment of predictors for the outcome and other predictors. | n.a. |
| Sample size | 8 | Explain how the study size was arrived at. | 7 |
| Missing data | 9 | Describe how missing data were handled (e.g., complete-case analysis, single imputation, multiple imputation) with details of any imputation method. | 7 |
| Statistical analysis methods | 10a | Describe how predictors were handled in the analyses. | 8-11 |
|  | 10b | Specify type of model, all model-building procedures (including any predictor selection), and method for internal validation. | 8-10 |
|  | 10d | Specify all measures used to assess model performance and, if relevant, to compare multiple models. | 9-11 |
| Risk groups | 11 | Provide details on how risk groups were created, if done. | n.a. |
| **Results** |  |  |  |
| Participants | 13a | Describe the flow of participants through the study, including the number of participants with and without the outcome and, if applicable, a summary of the follow-up time. A diagram may be helpful. | 7, 12 |
|  | 13b | Describe the characteristics of the participants (basic demographics, clinical features, available predictors), including the number of participants with missing data for predictors and outcome. | 12 |
| Model development | 14a | Specify the number of participants and outcome events in each analysis. | 12 |
|  | 14b | If done, report the unadjusted association between each candidate predictor and outcome. | n.a. |
| Model specification | 15a | Present the full prediction model to allow predictions for individuals (i.e., all regression coefficients, and model intercept or baseline survival at a given time point). | 14-15 |
|  | 15b | Explain how to use the prediction model. | n.a. |
| Model performance | 16 | Report performance measures (with CIs) for the prediction model. | 16 |
| **Discussion** |  |  |  |
| Limitations | 18 | Discuss any limitations of the study (such as nonrepresentative sample, few events per predictor, missing data). | 21-22 |
| Interpretation | 19b | Give an overall interpretation of the results, considering objectives, limitations, and results from similar studies, and other relevant evidence. | 19 |
| Implications | 20 | Discuss the potential clinical use of the model and implications for future research. | 21-22 |
| **Other information** |  |  |  |
| Supplementary information | 21 | Provide information about the availability of supplementary resources, such as study protocol, Web calculator, and data sets. | 27ff |
| Funding | 22 | Give the source of funding and the role of the funders for the present study. | n.a. |

### S4: STROBE Statement

STROBE Statement—Checklist of items that should be included in reports of *cohort studies*

S4: STROBE Statement STROBE Statement—Checklist of items that should be included in reports of cohort studies
|  | Item No | Recommendation | Page No |
| --- | --- | --- | --- |
| Title and abstract | 1 | (a) Indicate the study's design with a commonly used term in the title or the abstract | - |
|  |  | (b) Provide in the abstract an informative and balanced summary of what was done and what was found | 2 |
| Introduction |  |  |  |
| Background/rationale | 2 | Explain the scientific background and rationale for the investigation being reported | 4 |
| Objectives | 3 | State specific objectives, including any prespecified hypotheses | 5 |
| Methods |  |  |  |
| Study design | 4 | Present key elements of study design early in the paper | 6 |
| Setting | 5 | Describe the setting, locations, and relevant dates, including periods of recruitment, exposure, follow-up, and data collection | 6 |
| Participants | 6 | (a) Give the eligibility criteria, and the sources and methods of selection of participants. Describe methods of follow-up | 6 |
|  |  | (b) For matched studies, give matching criteria and number of exposed and unexposed | n.a. |
| Variables | 7 | Clearly define all outcomes, exposures, predictors, potential confounders, and effect modifiers. Give diagnostic criteria, if applicable | 13 |
| Data sources/measurement | 8* | For each variable of interest, give sources of data and details of methods of assessment (measurement). Describe comparability of assessment methods if there is more than one group | 6, 7 |
| Bias | 9 | Describe any efforts to address potential sources of bias | 10,11,17 |
| Study size | 10 | Explain how the study size was arrived at | 6,7 |
| Quantitative variables | 11 | Explain how quantitative variables were handled in the analyses. If applicable, describe which groupings were chosen and why | n.a. |
| Statistical methods | 12 | (a) Describe all statistical methods, including those used to control for confounding | 9-11 |
|  |  | (b) Describe any methods used to examine subgroups and interactions | n.a. |
|  |  | (c) Explain how missing data were addressed | 7 |
|  |  | (d) If applicable, explain how loss to follow-up was addressed | n.a. |
|  |  | (e) Describe any sensitivity analyses | n.a. |
| Results |  |  |  |
| Participants | 13* | (a) Report numbers of individuals at each stage of study—eg numbers potentially eligible, examined for eligibility, confirmed eligible, included in the study, completing follow-up, and analyzed | 12 |
|  |  | (b) Give reasons for non-participation at each stage | 12 |
|  |  | (c) Consider use of a flow diagram | 7 |
| **Descriptive data** | 14* | (a) Give characteristics of study participants (eg demographic, clinical, social) and information on exposures and potential confounders | 12 |
|  |  | (b) Indicate number of participants with missing data for each variable of interest | 12 |
|  |  | (c) Summarize follow-up time (eg, average and total amount) | n.a. |
| **Outcome data** | 15* | Report numbers of outcome events or summary measures over time | 12-13 |
| **Main results** | 16 | (a) Give unadjusted estimates and, if applicable, confounder-adjusted estimates and their precision (eg, 95% confidence interval). Make clear which confounders were adjusted for and why they were included | 16 |
|  |  | (b) Report category boundaries when continuous variables were categorized | n.a. |
|  |  | (c) If relevant, consider translating estimates of relative risk into absolute risk for a meaningful time period | n.a. |
| **Other analyses** | 17 | Report other analyses done—eg analyses of subgroups and interactions, and sensitivity analyses | 17-18 |
| **Discussion** |  |  |  |
| **Key results** | 18 | Summarize key results with reference to study objectives | 19 |
| **Limitations** | 19 | Discuss limitations of the study, taking into account sources of potential bias or imprecision. Discuss both direction and magnitude of any potential bias | 21-22 |
| **Interpretation** | 20 | Give a cautious overall interpretation of results considering objectives, limitations, multiplicity of analyses, results from similar studies, and other relevant evidence | 19-21 |
| **Generalisability** | 21 | Discuss the generalisability (external validity) of the study results | 22 |
| **Other information** |  |  |  |
| **Funding** | 22 | Give the source of funding and the role of the funders for the present study and, if applicable, for the original study on which the present article is based | n.a. |

